# Associations between white matter hyperintensity burden, cerebral blood flow and transit time in small vessel disease: an updated meta-analysis

**DOI:** 10.1101/2020.10.06.20207373

**Authors:** Catriona R Stewart, Michael S Stringer, Yulu Shi, Michael J Thrippleton, Joanna M Wardlaw

**Author notes:** Correspondence: Prof J M Wardlaw, Centre for Clinical Brain Sciences, UK Dementia Research Institute Centre, University of Edinburgh, 49 Little France Crescent, Edinburgh, EH16 4SB.

## Abstract

Cerebral small vessel disease is a major contributor to stroke and dementia, characterised by white matter hyperintensities (WMH) on neuroimaging. WMH are associated with reduced cerebral blood flow (CBF) cross-sectionally, though longitudinal associations remain unclear. We updated a 2016 meta-analysis, identifying 30 studies, 27 cross-sectional (*n*=2956) and 3 longitudinal (*n*=440), published since 2016. Cross-sectionally, we meta-analysed 10 new studies with 24 previously reported studies, total 34 (*n*=2180), finding lower CBF to be associated with worse WMH burden (mean global CBF: standardised mean difference: −0.45, 95% confidence interval (CI): −0.64,-0.27). Longitudinally, the association of baseline CBF with WMH progression varied: the largest study (5 years, *n*=252) found no associations, while another small study (4.5 years, *n=*52) found that low CBF in the periventricular WMH penumbra predicted WMH progression. We could not meta-analyse longitudinal studies due to different statistical and methodological approaches. We found lower CBF within WMH compared to normal appearing white matter (novel meta-analysis; 5 cross-sectional studies; *n*=295; SMD: −1.51, 95% CI: −1.94,-1.07). These findings highlight that relationships between resting CBF and WMH are complex and that more longitudinal studies analysing regional CBF and subsequent WMH change are required to determine the role of CBF in small vessel disease progression.

## Introduction

Cerebral small vessel disease (SVD) develops from pathological changes in the blood vessels supplying the brain and is an underlying cause of about 20% of all strokes and the most substantial contributor to vascular dementia.^1^ Advances in neuroimaging have been key in identifying and elucidating common features of SVD, including white matter hyperintensities (WMH), although much remains unknown about the underlying pathological mechanisms and there are currently no effective treatments. Vascular changes may occur before WMH are detectable, and are thought to contribute to WMH development.^2^ However, whether CBF reductions precede or follow WMH progression remains controversial, as hypoperfusion could be a consequence of WMH rather than the cause.^3^ WMH can be reversible, therefore establishing initial processes involved in WMH development may help identify targets to prevent SVD progression and related pathology.^4, 5^

Our previous systematic review and meta-analysis of WMH and CBF found that more severe WMH burden was associated with lower CBF, although this association weakened when studies of patients with dementia and poor age matching of controls were excluded.^3^ Age, dementia and other vascular risk factors contribute to CBF reductions and can confound the relationship between WMH burden and CBF.^3^ Additionally, there were few data on sub-regional brain tissue CBF and longitudinal WMH changes, as most studies only examined whole brain and/or cortical CBF.

Since 2016, several studies have been published that provide data on regional or tissue-specific CBF in WMH and normal appearing white matter (NAWM) cross-sectionally,^6-10^ as well as longitudinal studies. Therefore, we updated our previous systematic review and meta-analysis^3^ to identify whether these gaps in the literature and previous inconsistencies have been resolved.

## Materials and methods

We updated the systematic review by Shi *et al*. (2016),^3^ covering from 1946 until December 2015, by conducting a literature search of MEDLINE and EMBASE from 1^st^ January 2016 up to 1^st^ February 2020, using OVID. The search strategy and methodological approach including quality assessment and data extraction were as published by Shi *et al*. (2016),^3^ combining exploded search terms relating to *small vessel disease* and *cerebral blood flow* with AND (Supplementary Figure 1). We identified additional records by hand-searching, from January 2016 to February 2020, the *Journal of Cerebral Blood Flow and Metabolism* and within the subject of Cerebrovascular Disease and Stroke in *Stroke*.

### Eligibility Criteria

We included longitudinal and cross-sectional studies investigating associations between CBF or transit time metrics, including arterial transit time (ATT) and mean transit time (MTT), and WMH in patients with SVD, as well as studies measuring CBF velocity (CBFv) using ultrasound Doppler. We included studies using magnetic resonance imaging (MRI), including with phase contrast, arterial spin labelling (ASL) or dynamic susceptibility contrast, positron emission tomography (PET), single-photon computed tomography and computed tomography (CT) perfusion assessment. We excluded studies in paediatric or animal populations, duplicates, conference abstracts and cross-sectional studies lacking analysable data on CBF.

### Data Extraction and Analysis

We screened all potentially eligible publications and extracted standardised data from studies meeting the eligibility criteria. We extracted data on the study population cohort, the type of study, SVD characteristics, measurements, and units of CBF reported. Where available, we extracted data on separate regional CBF measurements for WMH, defined using MRI or CT, and NAWM, as this was previously identified as a key gap in the literature.^3^ Where reported we also extracted data on associations between ATT or MTT and WMH. We assessed the quality of each included study using a checklist based on the Strengthening the Reporting of the Observational studies in Epidemiology (STROBE) criteria.^11^

We extracted mean and standard deviation (SD) CBF values from disease and control groups or according to WMH burden, where available from text, tables, graphs or, if necessary, supplemental material for cross-sectional studies. We recorded associations with WMH burden either alone or as a component of total SVD burden, such as the ordinal SVD score which includes a point for central or cerebral atrophy, lacunes and white matter hyperintensities when rated from CT scans,^12^ with cerebral microbleeds also included for MRI.^13^ We included cross-sectional studies with only qualitative data or association analysis on CBF and SVD in the review but not the meta-analysis. We also extracted association analysis data from studies which performed association analysis in addition to providing quantitative data. We extracted information from longitudinal studies on time to follow-up, baseline and follow-up measurements of CBF and WMH, and their associations. Where unavailable in the published materials, we contacted authors to request unpublished data on baseline and follow-up CBF and/or WMH volume.

### Meta-analysis

We included all studies reporting mean and SD values for CBF in patient groups by WMH burden in the cross-sectional meta-analysis. In studies with more than two patient groups of WMH severity, we took mean values to allow for a comparison between patients with negative-to-mild and moderate-to-severe WMH burden following a previously described procedure.^3^ We extracted CBF values for grey and white matter in various brain regions, as reported by individual studies. We calculated standardised mean differences (SMDs) and 95% confidence intervals (CIs) with a random effects model and analysed new data along with the data reported in our previous review.^3^ We also extracted CBF values where available from WMH and NAWM to perform a novel meta-analysis on regional variability in CBF in different white matter tissues. Where studies reported CBF values separately for periventricular and deep WMH, we took mean values to give a measurement of CBF in WMH. We used Cochrane Collaboration’s Review Manager (Revman version 5.4) to perform the meta-analyses.

## Results

We identified 783 articles published between January 2016 and February 2020 from the literature search, after removing duplicates (Figure 1). Of these, we identified 149 of potential relevance and screened full texts to assess eligibility, 119 were excluded primarily due to lack of analysable data (45), no data on the association between WMH and CBF (38) and conference abstracts (33). Overall, we included 30 studies for qualitative analysis, of which 15 studies provided data for meta-analysis (Figure 1). Of these 15, 10 cross-sectional studies were meta-analysed with data previously extracted from 24 cross-sectional studies published between 1946 and 2015,^3^ while 5 studies were included in the novel meta-analysis of regional variability in CBF within WMH and NAWM.

**Figure 1.**
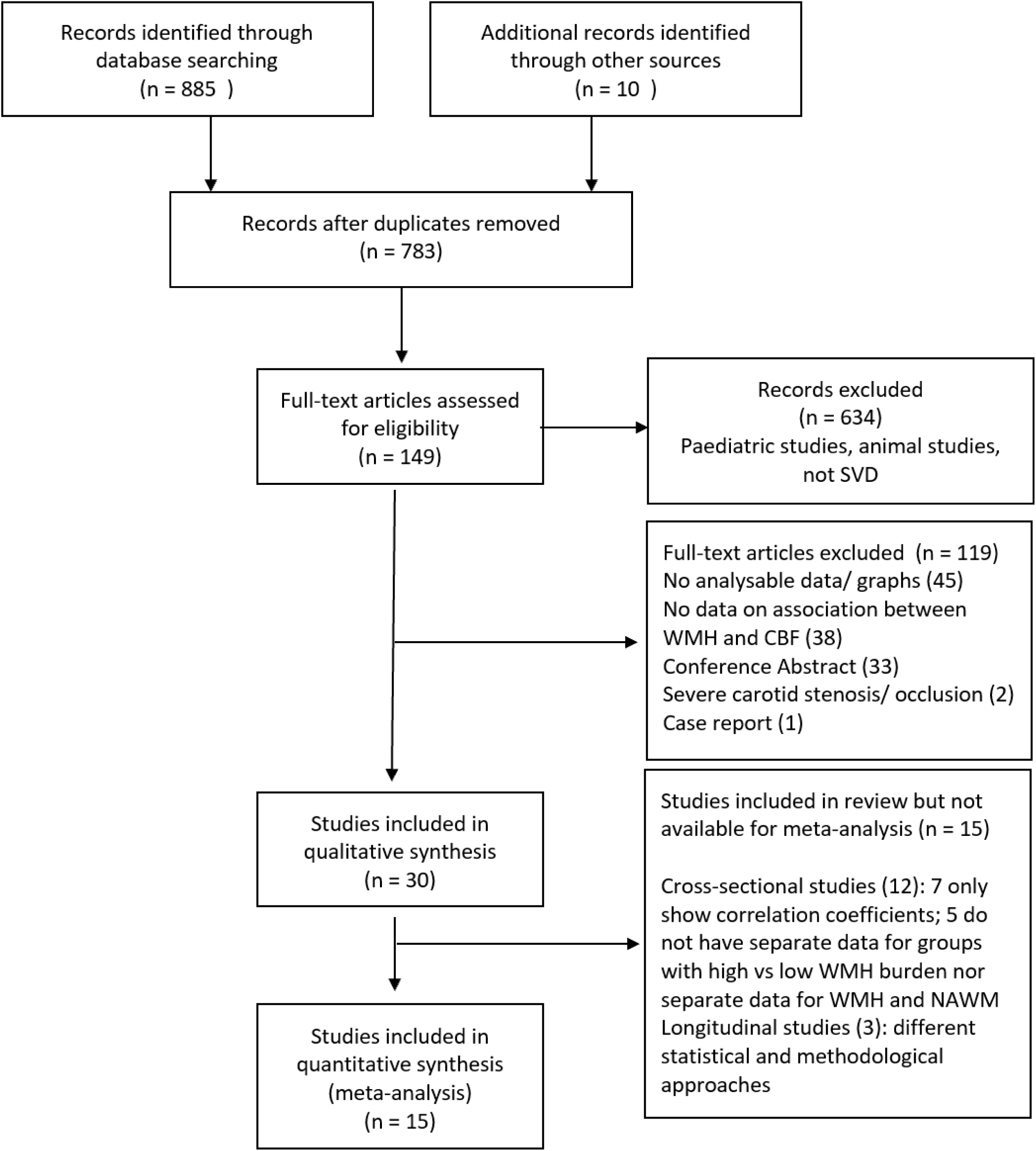
PRISMA flow diagram of literature search.

Some publications used the same study population cohort: Promjunyakul et al.^14, 15^ from the Layton Ageing and Alzheimer’s Disease Centre longitudinal ageing study and Turk et al.^16, 17^ with the same cohort of patients with ischaemic leukoaraiosis (ILA) and age, sex and risk-factor matched controls without ILA. We ensured that each participant was only included once in any single analysis.

### Characteristics of included studies

We extracted data from 27 new cross-sectional and 3 new longitudinal studies. All the new studies investigated patients or controls with varying degrees of WMH burden. The characteristics of these 30 studies are described in Table 1. The average study quality for all identified studies according to the STROBE criteria was 6.5/9 (Supplementary Figure 2). Main reasons for lower overall study quality scores included studies not reporting the number of participants who dropped-out (11/30), number and expertise of WMH assessors (13/30) and a lack of blinding to imaging or clinical data (14/30).

**Table 1.**
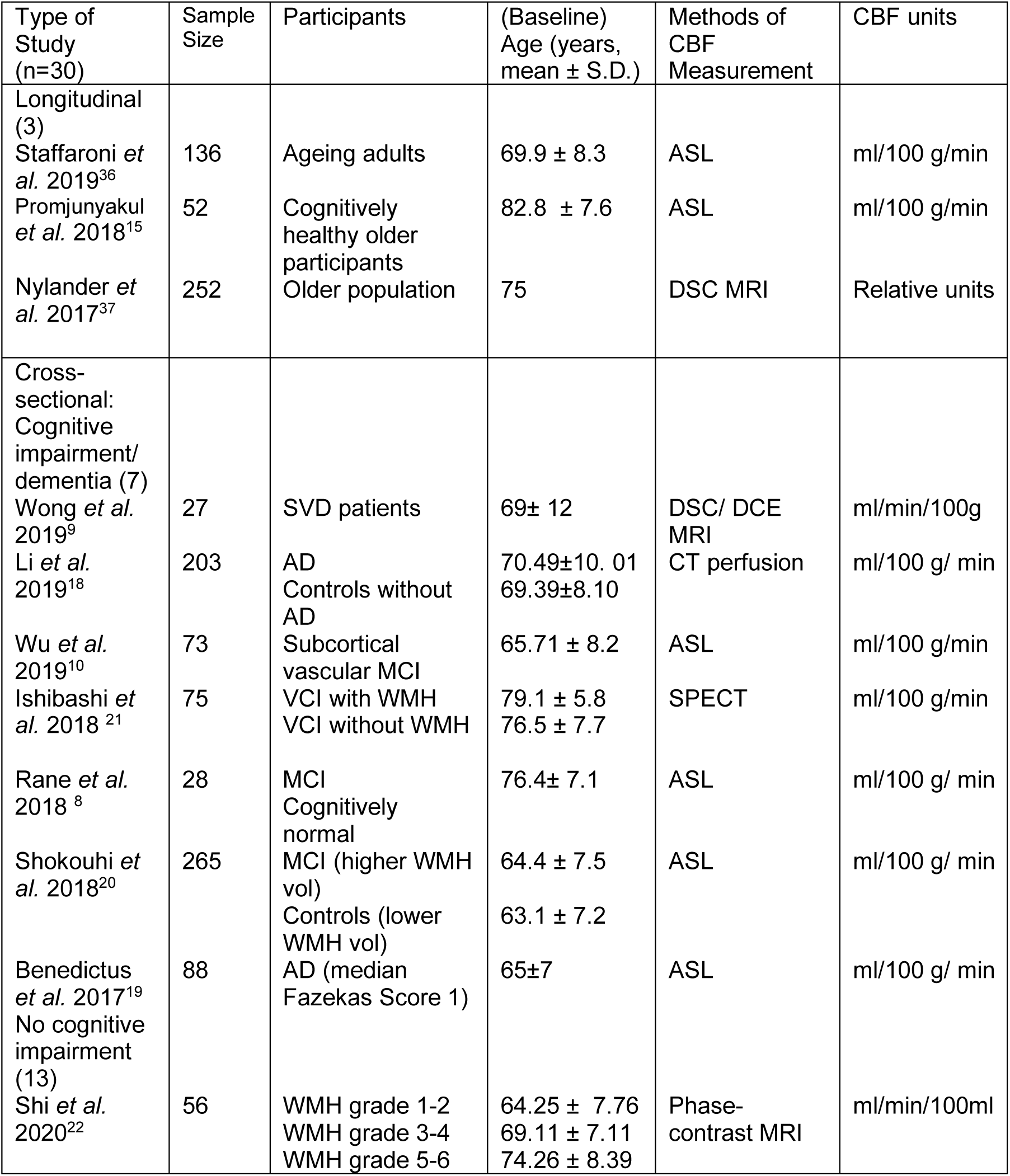
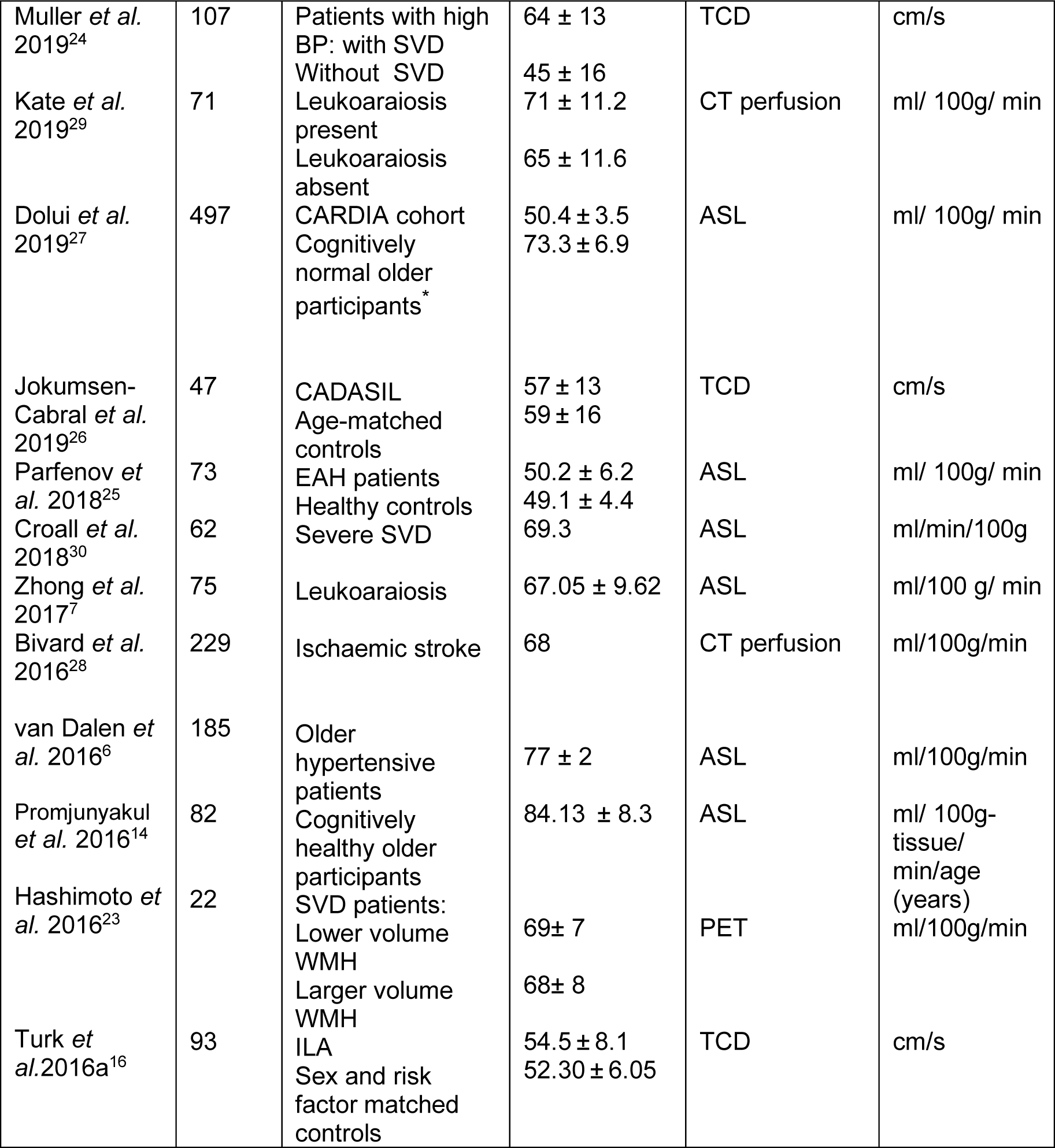
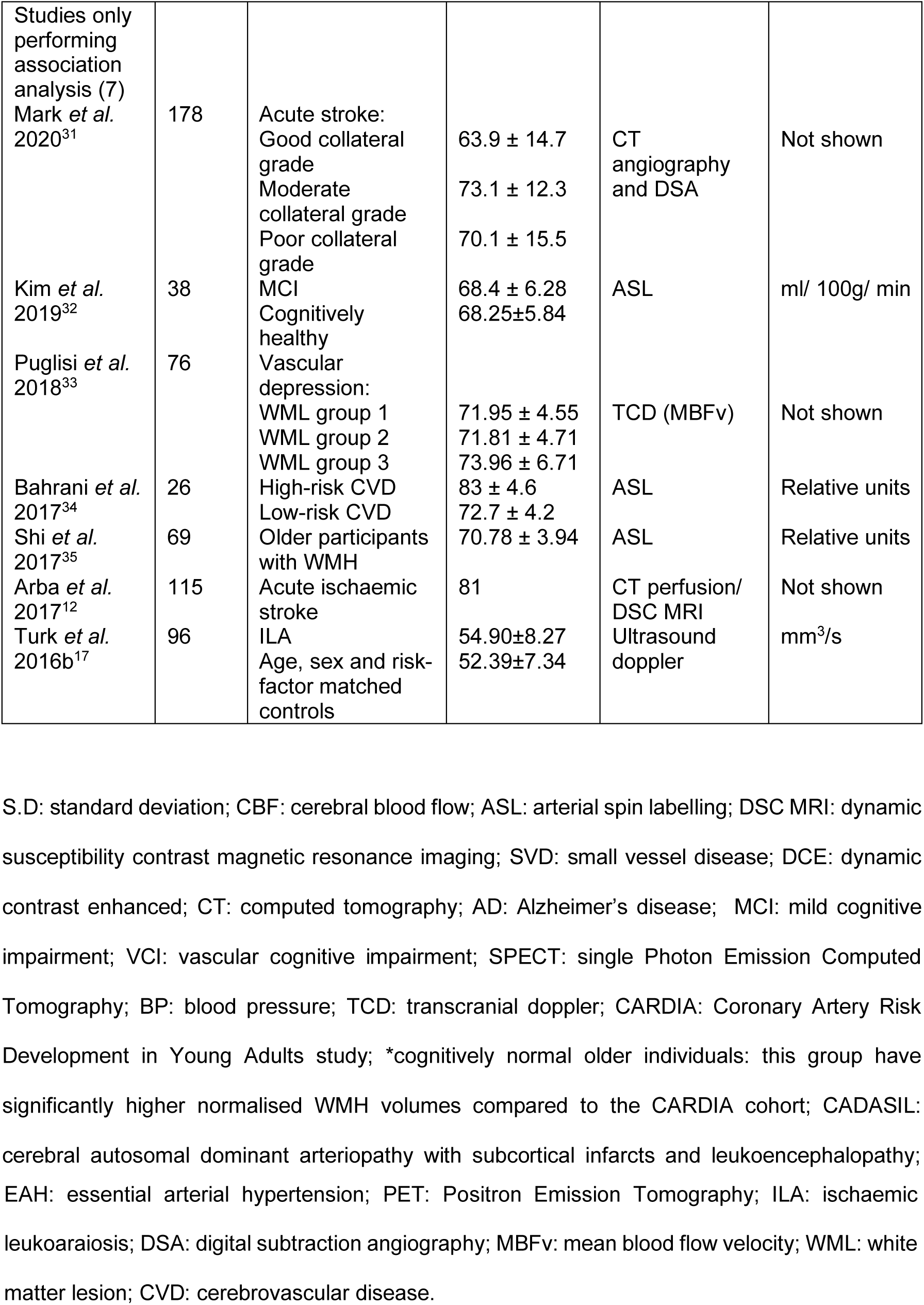
Characteristics of all studies included in this updated systematic review and meta-analysis.

Seven of the 27 newly identified cross-sectional studies investigated patients with cognitive impairment or dementia; including Alzheimer’s disease (AD),^18, 19^ mild cognitive impairment (MCI)^8, 10, 20^ and vascular cognitive impairment (VCI).^9, 21^

Thirteen of the 27 added cross-sectional studies investigated patients without cognitive impairment including patients with varying degrees of WMH burden,^7, 16, 22, 23^ hypertensive patients with and without SVD,^6, 24, 25^ individuals with Cerebral Autosomal Dominant Arteriopathy with Sub-cortical Infarcts and Leukoencephalopathy (CADASIL),^26^ cognitively normal older subjects^14, 27^ and patients with ischaemic stroke.^28^ Two of these 13 studies investigated the effect of blood pressure reduction on CBF in SVD, from which only baseline data on CBF and WMH burden, prior to any treatment, were extracted.^29, 30^

Seven of the 27 new cross-sectional studies reported only the results of association analyses, and did not provide quantitative data for CBF or WMH measurements. Patient groups in these studies included patients with acute stroke,^12, 31^ MCI,^32^ vascular depression^33^, individuals at varying risk of cerebrovascular disease^34^ and older individuals with WMH.^17, 35^

The three newly identified longitudinal studies (*n*=440) investigated baseline CBF and WMH progression in older individuals with WMH (Table 1) over follow-up periods ranging from two^36^ to five^37^ years. While 2 studies investigated associations between baseline WMH volume and change in either subregional^15^ or global^37^ CBF, one study considered associations between change in WMH volume and change in cortical grey matter CBF.^36^

Fifteen studies used ASL^6-8, 10, 14, 15, 19, 20, 25, 27, 30, 32, 34-36^, 3 used CT perfusion^18, 28, 29^, 2 used dynamic susceptibility contrast (DSC) MRI^9, 37^, one study each used SPECT^21^, phase contrast MRI^22^, CT angiography/digital subtraction angiography,^31^ PET,^23^ and a combination of CT and DSC MRI^12^. Five studies used transcranial Doppler ultrasound^16, 17, 24, 26, 33^.

### Cross-sectional CBF and WMH association

We extracted association analysis data from 11 of the new cross-sectional studies overall, including the 7 studies which reported association analyses without providing quantitative data and 4 studies which performed association analyses in addition to providing quantitative data.^6, 18, 27, 28^ Only 5 of these 11 studies adjusted for key vascular risk factors, including hypertension and smoking.^12, 17, 31, 33, 34^

### Cross-sectional analysis of associations between global or regional CBF and WMH

Four of the new studies analysed associations between global or regional CBF and WMH burden (Table 2). All of these studies found that patients with a worse WMH burden had lower CBF than those with a less severe WMH burden;^18, 28, 31, 32^ one study showed that worse PVWMH, DWMH and whole-brain WMH each showed a trend to be associated with lower CBF both in patients with AD and controls without AD (*n*=203).^18^ Worse WMH were associated with poorer cerebral collateral circulation, defined as cortical CBF at the outer surface of the brain, in one study which investigated patients with acute stroke (*n*=178).^31^

**Table 2.** Association analysis of a.) global or regional CBF with WMH; b.) mean blood flow velocity with WMH and c.) transit time with SVD burden.

| Study | Sample size | Statistical Method | Results Variables | Coefficients | P values | Adjusted for other variables |
| --- | --- | --- | --- | --- | --- | --- |
| a.) Global or regional CBF association with WMH |  |  |  |  |  |  |
| Bivard *et al.*<sup>28</sup> | 229 | Linear regression model | Hypoperfusion and likelihood of WMH | OR = 7.61 | **<0.001** | No |
| Kim *et al.*<sup>32</sup> | 38 | Linear regression analysis | Mean parietal lobe CBF and WMH volume | t(15) = -3 | **0.009** | Gender |
|  |  |  | Mean temporal lobe CBF and WMH volume | t(15)= -3.89 | **0.001** |  |
|  |  |  | Mean occipital lobe CBF and WMH volume | t(15)= -4.71 | **0.001** |  |
|  |  |  | Mean frontal lobe CBF and WMH volume | t(15)= -2.58 | **0.021** |  |
| Li *et al.*<sup>18</sup> | 203 | Spearman correlation analysis | Whole-brain WMH severity and CBF (AD patients) | r = -0.162 | 0.068 | Not shown |
|  |  |  | PVWMH severity and CBF (AD patients) | r = -0.219 | **0.013** |  |
|  |  |  | DWMH severity and CBF (AD patients) | r = -0.099 | 0.268 |  |
|  |  |  | Whole-brain WMH severity and CBF (controls) | r = -0.034 | 0.071 |  |
|  |  |  | PVWMH severity and CBF (controls) | r = -0.071 | 0.546 |  |
|  |  |  | DWMH severity and CBF (controls) | r = - 0.003 | 0.978 |  |
| Mark *et al.*<sup>31</sup> | 178 | Multivariate logistic regression analysis | PVWM Fazekas score and collateral flow<br>DWM Fazekas score and collateral flow<br>Total Fazekas score and collateral flow | NA<br>NA<br>NA | **<0.0001**<br>**<0.0001**<br>**<0.0001** | Age, gender, hypertension, hyperlipidemia coronary artery disease, atrial fibrillation, congestive heart failure and smoking history |
| b.) Mean blood flow velocity association with WMH |  |  |  |  |  |  |
| Turk *et al.* 2016b<sup>17</sup> | 96 | Spearman's correlation coefficient<br>Logistic regression | Correlation of average mean blood flow in ICA (mm3/s) with ILA<br>Association of average mean blood flow in ICA (mm3/s) with ILA | Spearman's p = -0.232<br>OR = 0.577 (0.368-0.902) | **0.025**<br>**0.016** | Age, gender and CVD risk factors |
| Puglisi *et al.*<sup>33</sup> | 76 | Spearman correlation | Correlation between MBFv in MCA and WMH severity | r = -0.34 | **0.002** | Demographic variables and clinical variables (including vascular risk factors) |
| c.) Transit time association with SVD burden |  |  |  |  |  |  |
| Arba *et al.*<sup>12</sup> | 115 | Logistic regression model | SVD score and mean transit time<br>SVD score and time to maximum flow<br>SVD score and time to peak<br>SVD score and arrival time fitted | OR = 2.80 (1.56-5.03)<br>OR = 2.36 (1.37-4.09)<br>OR = 1.52 (0.93-1.05)<br>OR = 3.59 (1.92-6.75) | **<0.05**<br>**<0.05**<br>N.S.<br>**<0.01** | Age, NIHSS, sex, hypertension and diabetes |

WMH: white matter hyperintensity; OR: odds ratio; CBF: cerebral blood flow; AD: Alzheimer's disease; PVWMH: periventricular white matter hyperintensity; DWMH: deep white matter
hyperintensity; PVWM: periventricular white matter; DWM: deep white matter; NA: not available; ICA: internal carotid artery; ILA: ischaemic leukoaraiosis; OR: odds ratio (95% confidence interval); CVD: cerebrovascular disease; MBFv: mean blood flow velocity; MCA: middle cerebral artery; SVD: small vessel disease; N.S: not significant; NIHSS: National Institutes of Health Stroke Scale.

Two of the new studies showed that lower mean blood flow velocity (MBFv) in the internal carotid artery and middle cerebral artery were associated with worse WMH burden in patients with ischaemic leukoaraiosis (*n*=96)^17^ and vascular depression (*n*=76)^33^ (Table 2).

### Cross-sectional association analysis comparing SVD burden and transit time

Arba *et al*. (2017)^12^ assessed SVD burden on CT or MRI, using an SVD score,^12^ and four transit time metrics (MTT, time to maximum flow, time to peak and arrival time fitted) derived from perfusion CT or DSC MRI, in the asymptomatic cerebral hemisphere of patients with acute ischaemic stroke (*n*=115). Worse SVD scores were associated with more prolonged transit times in white matter of the asymptomatic cerebral hemisphere (i.e. contralateral to the acute ischaemic stroke, Table 2). SVD burden was also associated with reduced CBF in the contralateral hemisphere.^12^

### Cross-sectional analysis of CBF in WMH and NAWM

Three recently published studies found that CBF was lower within WMH than in NAWM (Table 3). Higher WMH volume in older patients with hypertension (*n*=185)^6^ was associated with lower CBF within WMH. However, no clear relationship was seen between WMH volume and CBF in NAWM or grey matter (Table 3). Cognitively healthy older participants and healthy middle-aged participants (*n*=497) showed significantly lower CBF within WMH compared to normal appearing white matter.^27^ Similarly, in one small study (*n*=26), CBF was lower within DWMH and PVWMH compared to NAWM.^34^ In contrast, one exploratory study^35^ in healthy older participants (*n*=69) found in white matter regions where most patients had WMH; higher WMH burden correlated with higher grey matter perfusion in the opposite cerebral hemisphere.

**Table 3.** Association analysis comparing CBF within WMH and NAWM.

| Study | Sample size | Statistical Method | Results Variables Coefficients |  | P values | Adjusted for other variables |
| --- | --- | --- | --- | --- | --- | --- |
| Bahrani *et al.*<sup>34</sup> | 26 | Post-hoc paired comparisons | CBF in DWMH compared to NAWM | t = 5.7 | **<0.001** | Age, vascular risk group, total intracranial volume |
|  |  |  | CBF in PVWMH compared to NAWM | t = 11 | **<0.0001** |  |
|  |  |  | CBF in PVWMH compared to DWMH | t = 3.1 | **0.003** |  |
| van Dalen *et al.*<sup>6</sup> | 181 | Linear regression | WMH volume and WMH CBF | $\beta = -0.201$ | **0.029** | Total brain volume, age, antihypertensive use, brain parenchymal fraction, transit time |
| | | | WMH volume and NAWM CBF | $\beta = 0.175$ | 0.098 | |
| | | | WMH volume and GM CBF | $\beta = 0.175$ | 0.133 | |
| Dolui *et al.*<sup>27</sup> | 497 | Two-way repeated measures ANOVA | Mean CBF inside WML compared to outside (ADC cohort) | partial $\eta^2 = 0.75$ | **<0.0001** | No |
| | | | Mean CBF inside WML compared to outside (CARDIA cohort) | partial $\eta^2 = 0.59$ | **<0.0001** | |
| Shi *et al.*<sup>35</sup> | 69 | Pearson correlation | WMH ROI (R hemisphere) and CBF ROI (L hemisphere) | r = 0.42 | **0.00045** | Age, sex, education-adjusted MoCA score, total WMH volume |
|  |  |  | WMH ROI (L hemisphere) and CBF ROI | r = 0.43 | **0.00037** |  |
|  |  |  | (R hemisphere) |  |  |  |

CBF: cerebral blood flow; WMH: white matter hyperintensity; NAWM: normal appearing white matter; DWMH: deep white matter hyperintensity; PVWMH: periventricular white matter hyperintensity; GM: grey matter; $\beta$ : standardised estimate in linear regression; WML: white matter lesion; ADC: Alzheimer's Disease Centre (this is a longitudinal, prospective cohort and the individuals are cognitively healthy); CARDIA: Coronary Artery Risk Development in Young Adults study; ROI: region of interest; R: right; L: left MoCA: Montreal cognitive assessment.

### Sensitivity analyses of age, dementia and key vascular risk factors

We further refined the cross-sectional association analyses by removing studies investigating patients with dementia, studies without age-matching and studies which did not adjust for key vascular risk factors. The direction of effect in the remaining studies did not change: lower blood flow velocity^17, 33^ and prolonged perfusion CT/DSC transit time metrics^12^ remained associated with WMH, as well as lower CBF within WMH compared to NAWM.^34^ Only one cross-sectional study investigated WMH in association with global or regional CBF while adjusting for key vascular risk factors, showing worse WMH was associated with poorer collateral blood flow.^31^

### Longitudinal analyses of WMH and CBF

The largest of the three new longitudinal studies (Nylander *et al*. 2017)^37^ in a cohort of 406 randomly selected 75 year old participants at baseline and 252 at follow-up, found no significant association between baseline CBF with WMH progression at a five year follow-up when adjusting for baseline WMH and sex (Table 4). In 136 functionally normal older individuals, Staffaroni *et al*. (2019)^36^ found that longitudinal reductions in global CBF were associated with increasing WMH burden, adjusting for age, sex and education. Promjunyakul *et al*. (2018)^15^, in 52 cognitively healthy older participants, found that lower baseline CBF within the PVWMH penumbra was associated with WMH growth at follow-up, while reduced white matter perfusion remained predictive of PVWMH but not DWMH progression, adjusting for age and sex. These 440 patients from the three recent longitudinal studies add to the 1079 patients in 4 studies included in the 2016 review.^3^ Summarising all 7 longitudinal studies (*n*=1519), the two largest studies (total *n*=827, 575 + 252) found low baseline CBF did not predict worsening WMH; one previous and one new study (*n*=390 + 136) found low CBF predicted increased PVWMH but not deep WMH progression or predicted global WMH; the two smallest studies (*n*=40 and 52) found low CBF in NAWM/penumbral tissue predated WMH growth at follow-up; and one prior study (*n*=77) found CBF increased in some regions and decreased in other regions as WMH progressed.

**Table 4.** Results from longitudinal studies.

|  |  | Nylander et al.<sup>37</sup> | Staffaroni et al.<sup>36</sup> | Promjunyakul et al.<sup>15</sup> |
| --- | --- | --- | --- | --- |
| Sample size |  | 252 | 136 | 52 |
| Follow-up time (years) |  | 5 | 2.3 | 4.5 |
| CBF | At baseline | NA | 27.07 ± 6.38* | WM CBF: 27.0 ± 6.6<br>GM CBF: 59.3 ± 10.0<br>PVWMH CBF: 14.09 ± 6.05<br>DWMH CBF: 16.12 ± 6.85 |
| CBF | At follow-up | NA | 24.47 ± 5.61* | NA |
| WMH (ml) | At baseline | 10.5 ± 5.2<br>(n=406) | 3.4 ± 3.8* | Mean PVWMH vol. = 10.3 ± 11.7<br>Mean DWMH vol. = 1.63 |
| WMH (ml) | At follow-up | 11.9 ± 5.7<br>(n=229) | 4.6 ± 4.8 * | Mean PVWMH vol. growth = 3.7 ± 6ml<br>Mean DWMH vol. growth = 0.4 ± 1 ml |
| Baseline CBF and baseline WMH volume | Coefficient | NA | $\beta$ = -0.08,<br>b = -0.01 | NA |
|  | P-value | N.S. | 0.358 |  |
| Baseline CBF and ΔWMH volume | Coefficient | NA | NA | NA |
|  | P-value | N.S. |  |  |
| Baseline WMH volume and ΔCBF | Coefficient | NA | NA | NA |
|  | P-value |  |  |  |
| ΔWMH volume and ΔCBF | Coefficient | NA | b = -0.02 | NA |
|  | P-value |  | 0.007 |  |
| Adjusted for other variables |  | Baseline value, sex | Age, sex, education | Age, sex |

CBF: cerebral blood flow; WM: white matter; GM: grey matter; PVWMH: periventricular white matter hyperintensity; DWMH: deep white matter hyperintensity; WMH: white matter hyperintensity; vol.: volume; standardized beta ( $\beta$ ) and unstandardized (b): estimates given for
linear regression models; bCBF: baseline CBF; bWMH: baseline WMH; $\Delta$ WMH: change in WMH; $\Delta$ CBF: change in CBF; NA: not available. \*Unpublished data received from author upon request.

Longitudinal meta-analysis was not possible due to insufficient data and different statistical and methodological approaches between studies. The two studies with follow-up CBF measures^36, 38^ carried out their statistics differently and therefore could not be combined.

### Cross-sectional meta-analysis of CBF and WMH burden

We included 34 cross-sectional studies (*n*=2180) in this updated meta-analysis, of which 24 had been previously identified.^3^ The 10 newly identified studies covered 4 brain regions, including mean global CBF,^18, 21, 22, 25^ mean basal ganglia blood flow,^23^ total grey matter^6, 20^ and centrum semiovale^23^ (Figure 2). We extracted data on CBF velocity, as measured from the middle cerebral artery (vMCA) ^16, 24, 26^ and included this under a new subgroup in the meta-analysis. We found no additional published data for 7 brain regions included in our previous review: four in grey matter (frontal, temporal, parietal and occipital) and three in white matter (total, frontal and occipital).

**Figure 2.**
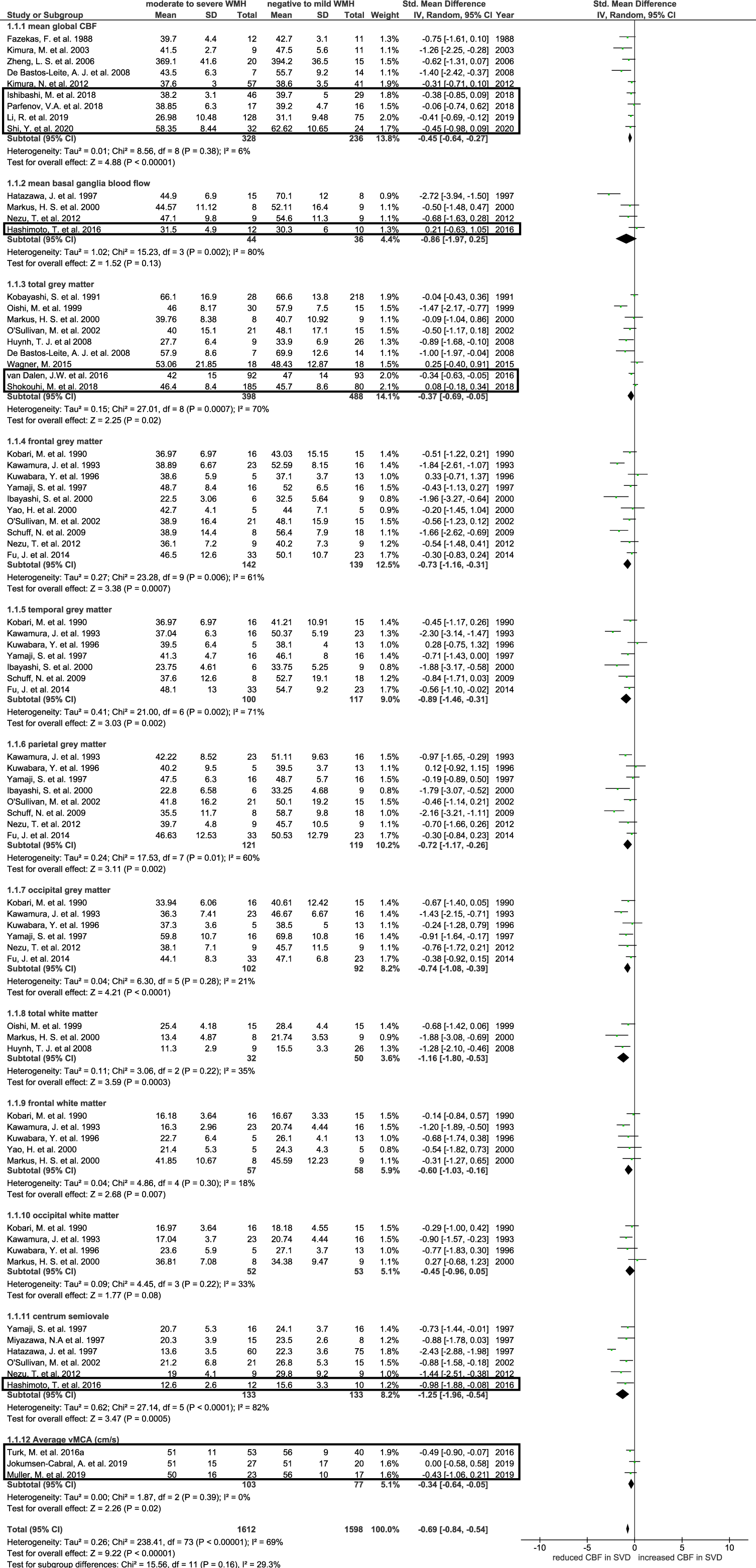
Forest plot showing standard mean differences in global, grey matter and white matter cerebral blood flow and in blood flow velocity in the middle cerebral artery. Cerebral blood flow was analysed in subgroups according to brain regions. Additional studies identified since our previous meta-analysis are outlined, all other data was originally published by Shi *et al*. (2016). CBF: cerebral blood flow; WMH: white matter hyperintensity; vMCA: blood flow velocity in middle cerebral artery.

Of the four regions where novel data was identified, patients with moderate-to-severe WMH burden had lower CBF than those with negative-to-mild WMH burden in three regions: global CBF (SMD: −0.45, 95% CI: −0.64, −0.27), total grey matter (SMD: −0.37, 95% CI: −0.69, −0.05) and centrum semiovale (SMD: −1.25, 95% CI: −1.96, −0.54) (Figure 2). In the basal ganglia there was no difference in CBF between patients with moderate-to-severe and negative-to-mild WMH burden (SMD: −0.86, 95% CI: −1.97, 0.25). Participants with worse WMH burden had lower blood flow velocities in the middle cerebral artery (SMD: −0.34, 95% CI: −0.64, −0.05).

### Sensitivity analyses of age and dementia

As with the previous review,^3^ the difference in CBF between patients with moderate-to-severe and negative-to-mild WMH burden attenuated and became non-significant upon removing studies investigating patients with dementia and further removing non-age matched studies from the updated meta-analysis, apart from mean global CBF and centrum semiovale CBF (Supplementary Figures 3 and 4). Mean middle cerebral artery blood flow velocity was not significantly different on comparing individuals with moderate-to-severe and negative-to-mild WMH burden after excluding non-age matched studies. Seven of the ten additional studies were included in the meta-analysis after removing studies which investigated patients with dementia and/or without age-matching; and the studies included in the mean basal ganglia and centrum semiovale categories did not differ, i.e. none of these studies included patients with dementia/without age-matching.

### Cross-sectional meta-analysis of CBF difference between WMH and NAWM

Additional to the analyses that were possible in 2016, we found sufficient data for meta-analysis in five cross-sectional studies reporting CBF values for both WMH and NAWM in subjects with moderate to severe WMH burden (*n* = 295). CBF was lower in WMH compared to NAWM (SMD:-1.51, 95% CI: −1.94, −1.07) (Figure 3).

**Figure 3.**
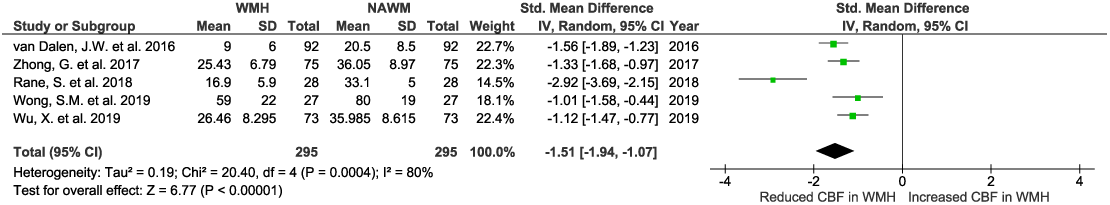
Forest plot of novel meta-analysis, showing standard mean differences in CBF in WMH compared to NAWM in subjects with moderate to severe WMH burden. WMH: white matter hyperintensity; CBF: cerebral blood flow; NAWM: normal appearing white matter.

## Discussion

We identified 30 new relevant publications since 2016, providing a total additional sample of 3,396. The newly identified cross-sectional studies reported lower CBF in most patients with higher WMH burden, supporting previous findings.^3^ In novel meta-analysis, we confirmed that CBF was also lower in WMH than in NAWM. Longitudinal findings were mixed, the largest studies (*n*=575+252) found baseline global CBF was not associated with WMH progression,^37^ as such there is no definite association between low CBF and increasing WMH long term. However, regional associations between lower CBF and WMH progression, notably in normal-appearing tissue surrounding PVWMH (*n*=390+52),^15, 38^ may exist.

Higher WMH burden was also associated with a lower blood flow velocity in the middle cerebral artery, which could be an important contributor to hypoperfusion.^17, 33^ One cross-sectional study showed that higher SVD burden was associated with prolonged MTT on CT and DSC.^12^

Only a minority (5/11) of cross-sectional studies and none of the longitudinal studies adjusted for cardiovascular risk factors. Cardiovascular risk factors can influence the development and rate of progression of SVD, while also affecting CBF via several mechanisms.^39^ Hypertension is an established risk factor for WMH that may affect CBF through vascular morphological changes and lumen narrowing, which can contribute to hypoperfusion.^40^ One cross-sectional study (*n*=73) found cerebral perfusion was lower in patients with hypertension, with or without WMH, than in healthy controls, though sample size was limited.^25^ Meanwhile diabetes has been linked to increased blood viscosity and a reduction in deformability of erythrocytes proportional to the degree of microvascular complications;^41^ white matter CBF is also lower in patients with type 2 diabetes than in controls.^42^ As such, controlling for cardiovascular risk factors is crucial in avoiding potential confounds.

### Strengths and weaknesses

Strengths of this work include the thorough meta-analysis of regional CBF variability by WMH burden, obtained through updating previous work.^3^ The sample size for studies included in the cross-sectional meta-analysis was enhanced (*n* = 2180, 64/study). We also screened non-English language publications, including a paper published in Russian.^25^ Limitations of this work include the lack of additional data in some brain subregions, though attempts were made to contact several authors to acquire unpublished data. However, this may also reflect a narrowing focus to key regions based on experimental hypotheses, excluding regions which are less reproducible. The meta-analysis of CBF variability in WMH and NAWM was limited by the small number of studies available (5), although the average sample size was reasonable (*n* = 295, mean 59/study). Several studies lacked control participants with little to no WMH burden and so could not be included. Longitudinal studies remain very limited and used heterogeneous methods, preventing direct comparison and longitudinal meta-analysis. Follow-up CBF measurements were only reported in 2 longitudinal studies;^36, 38^ thus interpretation of CBF changes over time in association with WMH progression is limited. However, an increasing use of non-contrast perfusion methods, particularly ASL, should make longitudinal imaging more viable. The role of other SVD markers, including lacunes and enlarged perivascular spaces, is also underexplored. Lastly, the accuracy of white matter and WMH perfusion measurements using ASL remains unclear, due to the lower perfusion level and signal-to-noise ratio relative to grey matter. However, there are on-going efforts to improve sensitivity of acquisition and analysis techniques.^43^ Additionally, none of the included ASL studies measured ATT, perhaps because ATT quantification requires a longer acquisition. ^44^

### Interpretation in relation to CBF and WMH pathology

WMH and NAWM CBF variability remains underexplored and more detailed longitudinal studies evaluating subregional changes in tissue CBF and WMH progression are needed. CBF reductions were more severe within WMH compared to NAWM in patients with moderate-to-severe WMH burden, suggesting WMH represent areas of focal perfusion deficits which are more severe than global hypoperfusion. However, associations with CBF may also be affected by other mechanisms implicated in WMH evolution.^45^ Two cross-sectional studies suggested that perfusion may be higher with higher WMH burden in normal-appearing white and some grey matter regions,^6, 35^ but were non-conclusive. Compensatory mechanisms may also affect regional perfusion, with areas of increased perfusion potentially counteracting deficiencies elsewhere.^46^ However, it is unclear whether compensatory perfusion changes would be sustained if they do occur, and could instead indicate dysfunctional blood flow regulation with worse WMH burden. Replication and more detailed subregional analyses of tissue changes in larger cohorts over time are required. Subtle regional differences in perfusion may also be masked within larger regions.^15^ The selection of regions of interest should therefore be considered carefully, and may favour finer grained regions, such as stratifying tissue by proximity to existing WMH^47^ or other pathological features.

### Implications for future research

In conclusion, this systematic review and meta-analysis showed that CBF was lower in most grey and white matter regions of the brain with higher WMH burden and lower in WMH than normal appearing white matter. Further studies investigating CBF along with ATT may help give a more comprehensive understanding of causal and consequential effects in WMH development and progression. Longitudinal studies exploring the effect of CBF changes on WMH burden remain limited. Further longitudinal studies are required to better understand the role of perfusion in WMH development and evolution. Future studies should also adjust for key vascular risk factors known to influence CBF, particularly hypertension, and, where relevant, baseline WMH burden.

## Data Availability

All data can be shared upon request.

## Acknowledgements

We would like to thank Tetiana Poliakova for translating the Russian article included in this review and Wenwen Li and Junfang Zhang for screening a Mandarin paper which did not satisfy the inclusion criteria. We thank Dr Francesca Chappell for providing advice on statistical analysis.

## Author Contributions

M.S.S. and J.M.W. conceived the idea of the study. Y.S. and J.M.W. designed the search strategy and provided data files from the original literature review. C.R.S. did the data search, extracted data and conducted the statistical analyses. M.S.S. cross-checked the data. C.R.S. drafted the report and designed the tables and figures. J.M.W. obtained funding and managed the project. All authors revised and approved the manuscript.

## Disclosure/ conflict of interest

The Authors declare that there is no conflict of interest.

## Funding

The authors disclosed receipt of the following financial support for the research, authorship and/or publication of this article: NHS Lothian R+D Department (MJT); the UK Dementia Research Institute which receives its funding from DRI Ltd, funded by the UK MRC (MSS, MJT, JMW); the Fondation Leducq Transatlantic Network of Excellence for the Study of Perivascular Spaces in Small Vessel Disease [reference number 16 CVD 05] (MSS); and European Union Horizon 2020 [project number 666881, SVDs@Target] (MSS).

## Supplemental Information

Please find all supplementary figures in the Supplemental Material document.

## Notes

### Competing Interest Statement

The authors have declared no competing interest.

### Author Declarations

N.A. as this is not a critical trial.

