## Supplemental Material for "Associations between white matter hyperintensity burden, cerebral blood flow and transit time in small vessel disease: an updated meta-analysis"

**Supplementary Figure 1.** Systematic review search strategy.

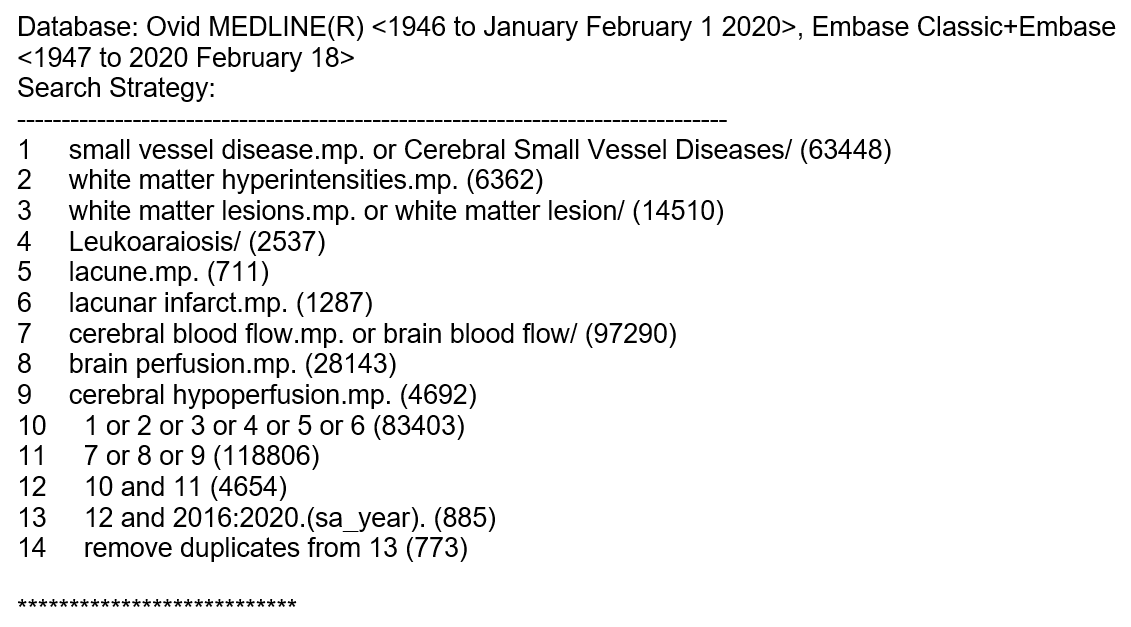

**Supplementary Figure 2.** Study quality according to the Strengthening the Reporting of the Observational studies in Epidemiology (STROBE) statement (Bailey *et al.* 2012).

| **Study** | **Overall Score** |
| --- | --- |
| Bivard *et al.* 2016 | 8 |
| Hashimoto *et al.* 2016 | 7 |
| Turk *et al.* 2016a | 5 |
| Turk *et al.* 2016b | 7 |
| van Dalen *et al.* 2016 | 8 |
| Promjunyakul *et al.* 2016 | 6 |
| Bahrani *et al.* 2017 | 6 |
| Arba *et al.* 2017 | 8 |
| Shi *et al.* 2017 | 6 |
| Zhong *et al.* 2017 | 7 |
| Benedictus *et al.* 2017 | 7 |
| Nylander *et al.* 2017 | 9 |
| Ishibashi *et al.* 2018 | 7 |
| Promjunyakul *et al.* 2018 | 5 |
| Rane *et al.* 2018 | 7 |
| Puglisi *et al.* 2018 | 6 |
| Croall *et al.* 2018 | 7 |
| Shokouhi *et al.* 2018 | 6 |
| Parfenov *et al.* 2018 | 4 |
| Jokumsen-Cabral *et al.* 2019 | 6 |
| Wong *et al.* 2019 | 7 |
| Kim *et al.* 2019 | 6 |
| Li *et al.* 2019 | 8 |
| Dolui *et al.* 2019 | 5 |
| Staffaroni *et al.* 2019 | 6 |
| Wu *et al.* 2019 | 5 |
| Muller *et al.* 2019 | 5 |
| Kate *et al.* 2019 | 6 |
| Mark *et al.* 2020 | 6 |
| Shi *et al.* 2020 | 9 |

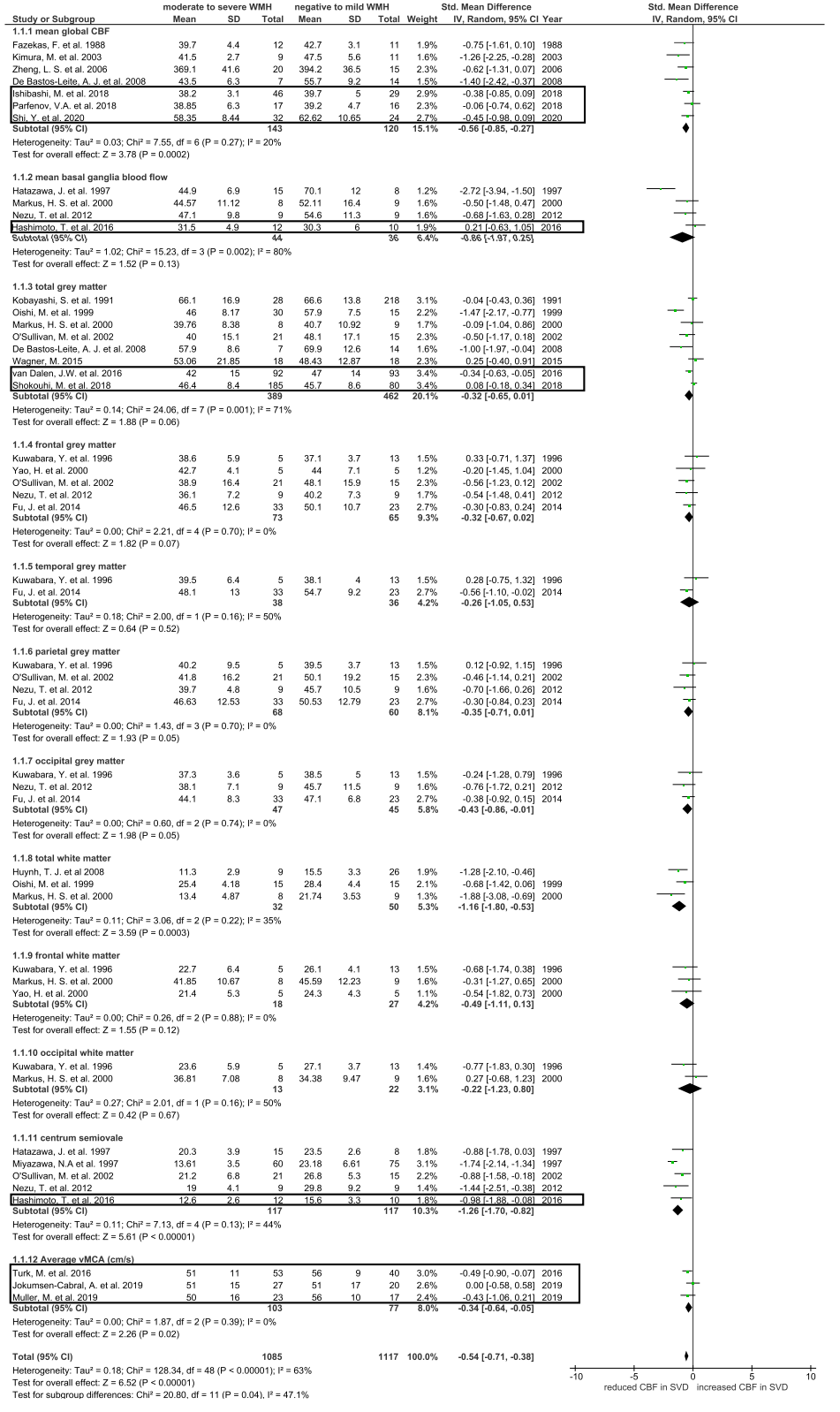
**Supplementary Figure 3.** Sensitivity analysis: forest plot showing standard mean differences in CBF in brain regions according to WMH burden, after excluding studies with dementia patients.

CBF: cerebral blood flow; WMH: white matter hyperintensity; vMCA: blood flow velocity in middle cerebral artery. Recent studies are outlined, all other studies were published in our previous review.

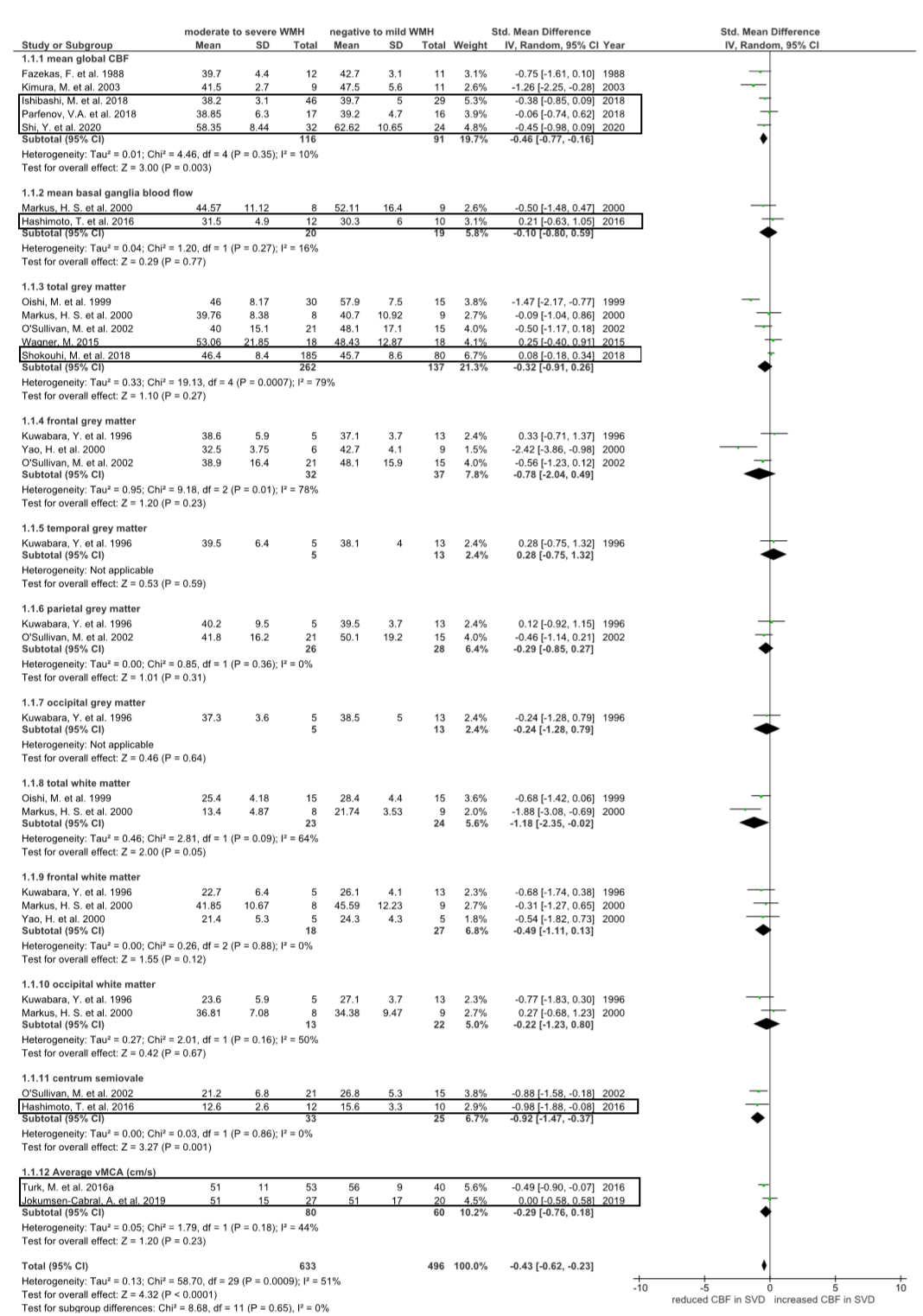
**Supplementary Figure 4.** Sensitivity analysis: forest plot showing standard mean differences in CBF in each brain region according to WMH burden, after excluding studies with dementia patients and studies without age-matching.

CBF: cerebral blood flow; WMH: white matter hyperintensity; vMCA: blood flow velocity in middle cerebral artery. Recent studies are outlined, all other studies were published in our previous review.
